# The SQUEEZE pilot trial: a trial to determine whether septic shock reversal is quicker in pediatric patients randomized to an early goal directed fluid-sparing strategy vs. usual care

**DOI:** 10.1101/2024.08.21.24312369

**Authors:** Melissa J. Parker, Karen Choong, Alison Fox-Robichaud, Patricia C. Liaw, Lehana Thabane

## Abstract

**Objective:** The overall objective of our research is to determine in children with septic shock whether use of a fluid-sparing strategy results in improved clinical outcomes without an increased risk of adverse events compared to usual care. The specific objective of this pilot randomized controlled trial was to evaluate the feasibility of a definitive multicenter trial to answer our research question.

**Design:** Pragmatic, 2-arm, parallel group, open label, prospective pilot randomized controlled trial including a nested biosample-based translational study.

**Setting:** Pediatric tertiary care centre

**Patients:** Children aged 29 days to <18 years of age presenting to the Emergency Department or admitted to an in-patient ward (including the PICU) with suspected or confirmed septic shock and a need for ongoing resuscitation.

**Interventions:** Fluid-sparing vs. usual care resuscitation strategy continued until shock reversal. The fluid-sparing intervention comprised instructions to restrict fluid bolus therapy in conjunction with early initiation and/or preferential use of vasoactive medication support as a strategy to spare fluid while targeting the hemodynamic goals specified in the American College of Critical Care Medicine Surviving Sepsis Guidelines. The usual care strategy did not limit use of fluid bolus therapy.

**Measurements and Main Results:** 53 were randomized to usual care (n=27) or fluid-sparing (n=26). Fifty-one participants were available for primary outcome analysis. Primary feasibility outcomes related to participant enrolment and protocol adherence. Enrolment rate was 1.8 (51/29); 95% confidence interval [CI]: 1.3-2.3 participants/month. Study procedures were implemented in 49/51 (96.1%), 95% CI: 86.5-99.5% participants within 1 hour of randomization in a median (quartile range [IQR]) of 8 (5, 15) minutes. The protocol required use of an exception to consent process and consent for ongoing participation was 48/51 (94.1%), 95% CI: 83.8-98.8%. There were no serious adverse events.

**Conclusions:** We concluded the large multicenter SQUEEZE Trial feasible to conduct. Trial Registration: ClinicalTrials.gov [NCT01973907]

## Introduction

Fluid resuscitation has long been a cornerstone of septic shock resuscitation.^1,2^ The 2009 Surviving Sepsis guidelines recommended early and aggressive fluid resuscitation, including successive fluid boluses of 20 mL/kg up to and over 60 mL/kg, followed by initiation of vasoactive medication support for ongoing shock.^1^ The Fluid Expansion as Supportive Therapy (FEAST) trial published in 2011 prompted significant interest in guideline recommendations for fluid bolus therapy after demonstrating an increased risk of mortality in children treated with bolus fluids.^3^ Emerging observational evidence also began to link fluid overload with an increased risk of morbidity and mortality in adults and children.^4–6^ This in conjunction with a lack of randomized controlled trial (RCT) evidence in high-income countries raised the important question of whether fluid boluses were helpful or harmful in children with septic shock with access to advanced critical care.^7^

We embarked on the SQUEEZE trial research program to answer this question. We also sought to leverage trial resources to investigate the prognostic value of plasma cell-free deoxyribonucleic acid (cfDNA) levels in children as this biomarker has been demonstrated to be an indicator of poor outcome in adult ICU patients with sepsis.^8,9^ When we began this work in 2013, it was clear that equipoise did not exist for randomizing children to no bolus vs fluid bolus therapy because isotonic fluid resuscitation remained the standard of care. The evidence for the optimal timing of initiation of vasoactive medications relative to fluid boluses for septic shock resuscitation was also unclear.^1^ We therefore planned to investigate a fluid-sparing strategy consisting of restriction of fluid bolus therapy vs usual care of liberal fluid resuscitation in conjunction with earlier initiation and preferential escalation of vasoactive medication(s) in the fluid-sparing group. We recognized that it was important to enable enrolment prior to pediatric intensive care unit (PICU) admission, that use of an exception to consent process would be required, and that many potential pitfalls could derail our trial. We determined a pilot RCT was needed to evaluate the feasibility of the SQUEEZE trial protocol and hypothesized that the multicenter SQUEEZE trial, including a nested biomarker-based translational study, was feasible to conduct.

## Materials and Methods

The trial design was a pragmatic, two-arm, open-label, prospective pilot RCT. Approval for single-centre study conduct was granted by the Hamilton Integrated Research Ethics Board (HIREB) on June 4, 2013 (Project ID: 13-295), with an amendment allowing addition of an external site. As a trial which enrolled participants experiencing an individual medical emergency, the research ethics board approved protocol included use of an exception to consent (deferred consent) process as supported by the Canadian Tri-council policy statement guidelines and the Declaration of Helsinki.^10^ A summary of protocol versions and amendments is provided (**Table S1**). The trial was prospectively registered on ClinicalTrials.gov on October 23, 2013 [NCT01973907] prior to enrolment of the first participant. We prepared the study protocol following the Standard Protocol Items: Recommendations for Interventional Trials (SPIRIT Guidelines).^11,12^ Our protocol was published in Trials where information including the SPIRIT checklist, World Health Organization Trial Registration Data Set, and the schedule of enrollment, interventions, and assessments can be accessed.^13^ An extended CONSORT checklist for pilot and feasibility trials is provided (**Appendix S1**).^14^

The study was promoted to pediatric emergency department (PED) and PICU staff physicians, nurses and clinical trainees who assisted with timely identification of potentially eligible patients. Patients were screened and enrolled 24 hours/day, 7 days per week by the SQUEEZE trial research assistant or one of the investigators. We enrolled children with suspected or confirmed septic shock and a need for ongoing resuscitation presenting from various locations within a pediatric tertiary care centre. This included children 29 days to <18 years of age presenting via the PED, inpatient wards (medical emergency team (MET) activations), and the PICU. A minimum of 40 mL/kg (2 L for children >50 kg) of isotonic fluid bolus therapy within the previous 6 hours, and ongoing signs of shock were required for inclusion. Full inclusion and exclusion criteria are previously published and presented below.^13^

SQUEEZE Pilot Trial Eligibility Criteria

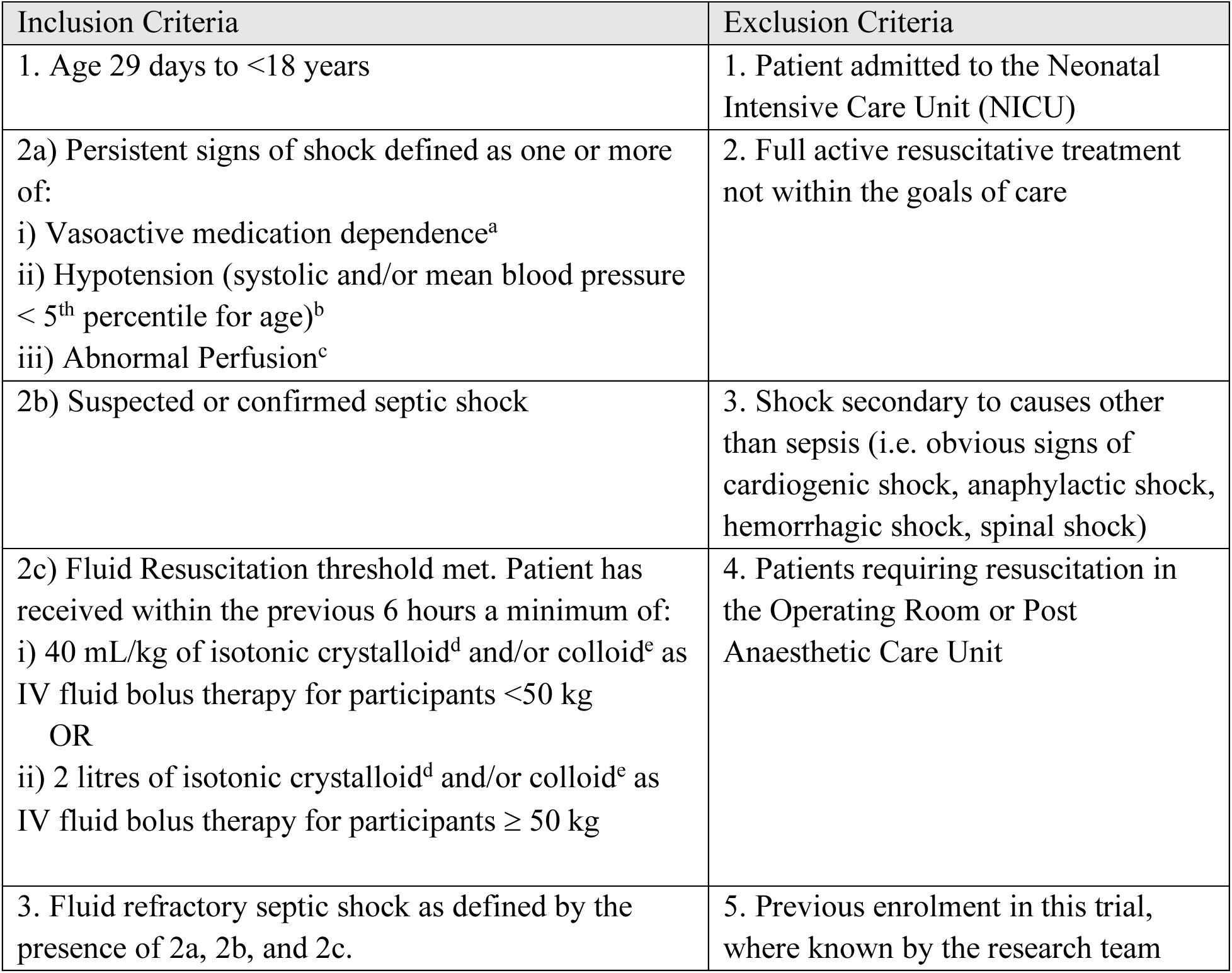
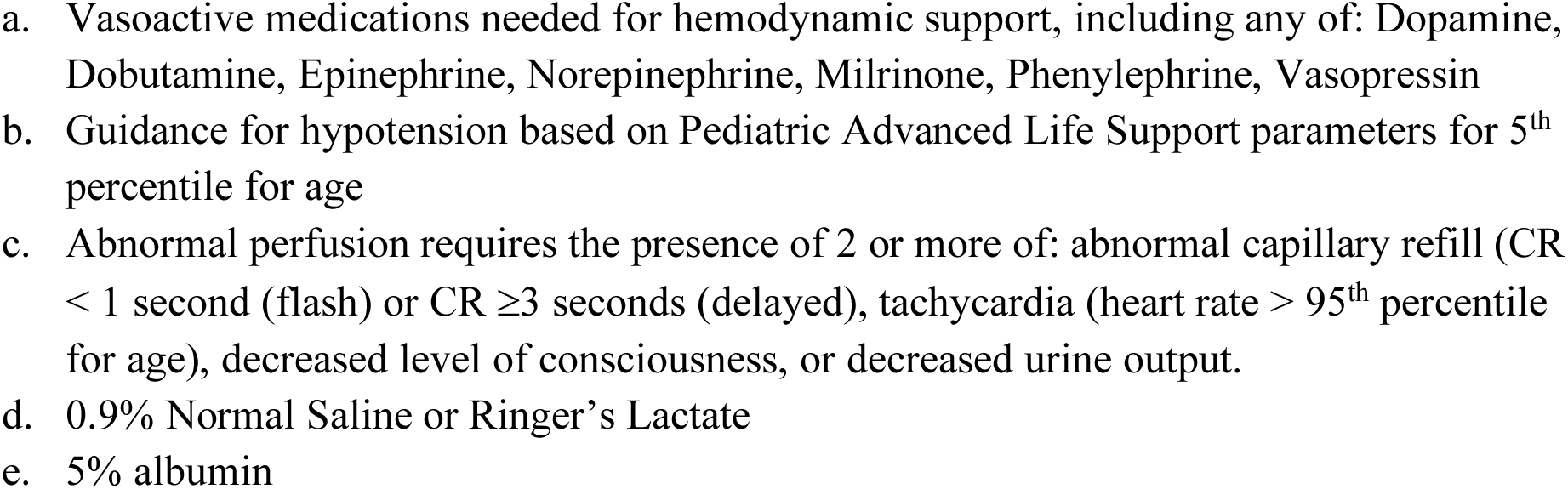

To identify eligible non-enrolled patients, we screened the daily PICU census and MET activation records. Eligible patients were randomized using a telephone accessible third party computer-based process. The allocation sequence was computer generated and prepared by the Biostatistics Unit using a schema of simple randomization with no stratification or blocking. The allocation sequence was kept secret from and inaccessible by the investigators.

Following randomization, the group assignment and initial instructions were communicated by the research team to the healthcare team managing the participant. Brief verbal instructions on how to implement the intervention were provided, including direction to obtain a SQUEEZE study package labelled with the group assignment. A detailed description of the intervention is presented elsewhere.^13^ In brief, the intervention had two tiers, with each tier providing instructions for vasoactive medication(s) use and fluid bolus therapy (**Figure 1**). For both groups, study packages included a copy of the ACCM hemodynamic goals and the surviving sepsis guideline to promote adherence to best practices for aspects of patient care not impacted by the intervention.^15^ A group specific one-page flow diagram provided instructions on how to implement the assigned treatment. Study signs noting the treatment assignment were placed on the medical record and in the patient’s room (at the head of the bed, tag on IV pump) as visible prompts for healthcare team members. For the fluid-sparing arm only, a fluid bolus record was provided to document provider justification for any administered fluid bolus(es). The assigned treatment was continued until shock was determined to be reversed. For the nested translational study (named SQUEEZE-D), blood samples were requested at two timepoints: Sample A (baseline, within 6 hours of randomization) and Sample B (24-48 hours following randomization). Samples were collected in a citrated plasma tube (CPT), spun, and stored within one hour of collection at -80 Celsius for later measurement of cell-free DNA (cfDNA).

**Figure 1.**
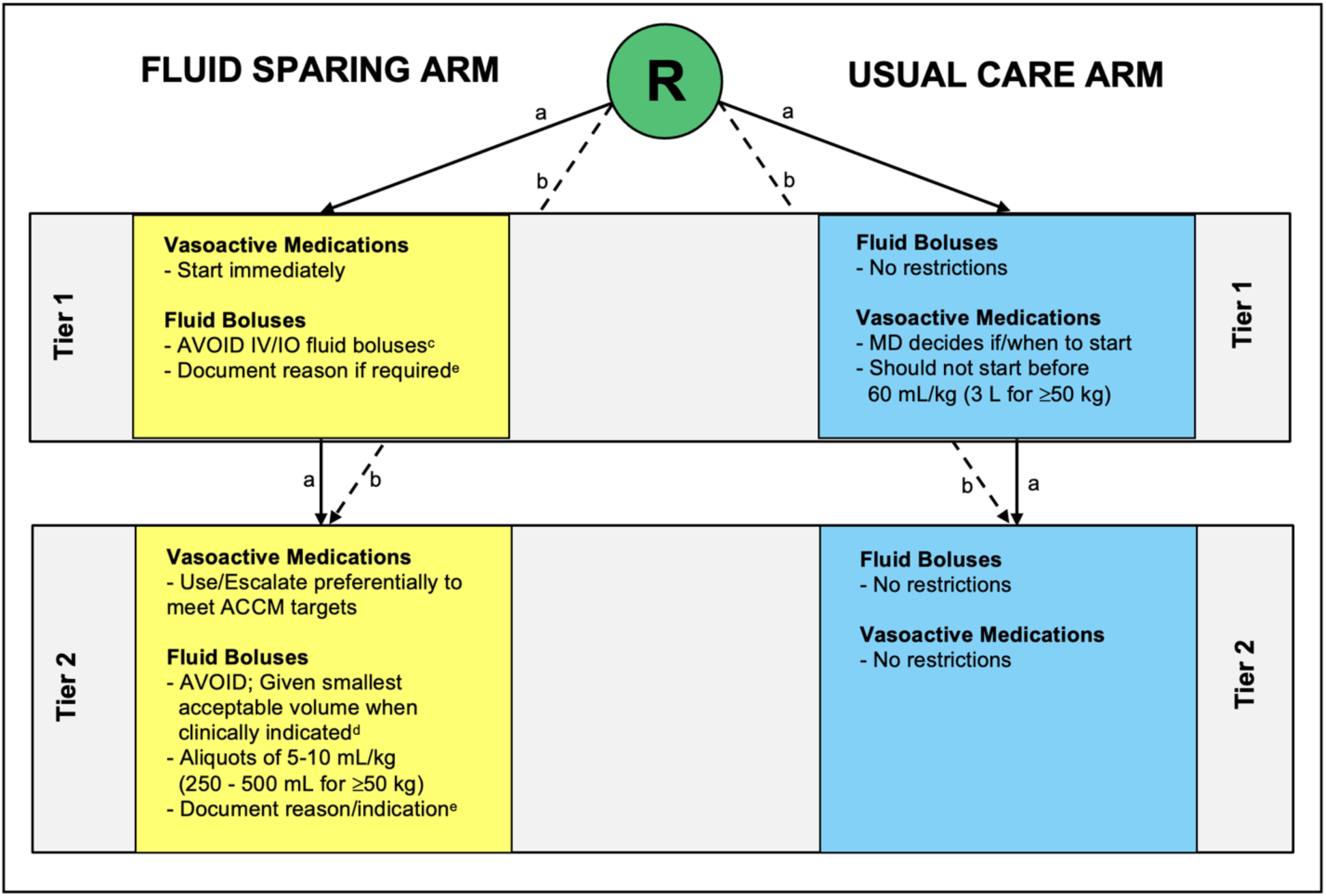
Fluid-sparing (intervention) and usual care (control) management strategies * This work is licensed under CC BY-NC 4.0 creativecommons.org/licenses/by-nc/4.0. Reproduced with permission from the original source file authored by Parker MJ, available at: http://hdl.handle.net/11375/29921. ^30^ **Legend for Figure 1.** a. Participants who are randomized prior to initiation of any vasoactive medication infusions begin the assigned allocation in **Tier 1**. Tier 1 contains instructions specific to the situation of a participant not being treated with vasoactive medications. A participant may never exit Tier 1. If vasoactive medications are commenced, the treatment instructions shift to those described for **Tier 2**. b. Participants who are randomized when already receiving a vasoactive medication infusion(s) begin the assigned allocation in **Tier 2**. Tier 1 is bypassed in such situations. c. Healthcare providers are requested to avoid giving any further fluid boluses. Valid reasons to give a small volume fluid bolus, as noted on the bedside study algorithm include: i) Clinically unacceptable delay in the ability to start a vasoactive medication infusion. (e.g. nursing staff need to prepare the infusion). ii) Documented intravascular hypovolemia. (based on clinician assessment) d. Healthcare providers are requested to avoid giving any further fluid boluses. Valid reason to give a small volume fluid bolus, as noted on the bedside study algorithm include: i) Documented intravascular hypovolemia. (based on clinician assessment) e. Healthcare provider justification for administration of any fluid bolus to a participant in the fluid-sparing group is to be documented on the fluid-sparing bolus record contained within the study package (fluid-sparing group only). Abbreviations R Randomization IV Intravenous IO Intraosseous mL millilitres kg kilograms MD medical doctor/physician

The purpose of conducting a pilot trial is to assess process, resource, management and scientific aspects of feasibility before embarking on a larger scale trial.^16^ The SQUEEZE pilot trial co-primary outcomes included, i) the ability to enroll participants, and ii) initiation of study procedures within 1 hour of randomization. We defined a priori the pass threshold for formal evaluation of protocol feasibility as the ability to enroll ≥2 participants per month (per site per month if a second site added). Due to the multifaceted nature of the intervention, the timing of initiation of study procedures was based on the time of initiation of the applicable treatment algorithm, which was recorded directly on the study package when implemented. Secondary outcomes included the appropriateness of eligibility criteria, the ability to collect clinical outcomes, and data related to study process, resource and management aspects of feasibility. This included feasibility of obtaining Ultrasonic Cardiac Output Monitor (USCOM^TM^) assessments of cardiac indices at specified timepoints. The primary feasibility outcome for the nested translational study (named SQUEEZE-D) was to determine the proportion of SQUEEZE participants for whom plasma levels of cfDNA could be described.^8,9^ SQUEEZE-D secondary outcomes included the availability of required samples, as well as study process, resource and management aspects of feasibility impacting specimen acquisition and testing.^13^

Demographic and clinical outcome data for participants were obtained from the medical record. Data were abstracted by trained research staff or one of the investigators and recorded on a paper data collection form for subsequent entry into the electronic REDCap Case Report Form (CRF).^17^ SQUEEZE-D specimen analysis results were retained in a secure file at the Thrombosis and Atherosclerosis Research Institute (TaARI).^13^ In accordance with Canadian research ethics policy, our protocol specified that data collected until consent decline would be retained to minimize risk of bias. Participants or their substitute decision makers (SDM) who declined the intervention were asked for permission to continue follow-up for data collection. Consent for SQUEEZE-D participation specifically as it pertained to use of biological specimens was documented within the same consent form.

We set the sample size for this pilot trial at 50 participants (25 per arm) as this is sufficient to evaluate feasibility.^16,18^ We did not base our sample size on a sample size calculation as this is not required for pilot studies. Considering both the fluid-sparing and usual care treatment strategies fell within the broad scope of Surviving Sepsis treatment recommendations, study participation was deemed minimal risk. Many clinical outcomes in the trial were categorized as adverse outcomes as is common in critical care research.^19^ Serious adverse events (SAEs) were defined in accordance with published REB guidance.^20^ We planned to report SAEs to the REB and to monitor these at the trial steering committee. We did not form a data safety and monitoring board (DSMB) for this pilot trial considering the minimal risk attributable to study participation and that study duration was too short for the DSMB process.^21^ There were no stopping rules or planned interim analysis.

The statistical analysis plan is described within our published pilot trial protocol and included descriptive summary measures for reporting of baseline characteristics and outcome variables (primary and secondary).^13^ This includes mean (SD) or median (Q1, Q3) for continuous variables and n (%) for categorical variables. We prespecified reporting of feasibility outcomes as descriptive estimates with 95% confidence intervals. The enrolment rate was estimated by dividing the number of participants enrolled by months of recruitment. Assessment of scientific aspects of feasibility included sample size calculations for the planned multicentre trial. We arranged for an independent statistician to perform the sample size calculation so that we could remain blinded to clinical outcome estimates. We planned to include pilot trial participants in the larger multicenter trial, if feasibility was confirmed, and as such we did not plan to analyze or report clinical outcomes beyond assessment for completeness. For SQUEEZE-D, the translational biosamples analysis results were similarly not further analyzed, with reporting planned with main trial results. We specified adherence to the Consolidated Standards of Reporting Trials (CONSORT) extension guidelines for reporting and analysis of pilot and feasibility RCTs and to have a statistician conduct the analyses using SAS (Cary, NC, USA).

## Results

The pilot trial was conducted at McMaster Children’s Hospital, a pediatric tertiary care centre in Hamilton, Canada. Participants were enrolled from January 6, 2014 to June 3, 2016 with follow-up to 90 days. The process of flow through the study is illustrated in **Figure 2** in accordance with CONSORT guidelines.^22,23^ At a single centre, there were 53 randomization occurrences in 52 unique individuals. This included two randomization errors, with one involving a previously enrolled participant. The analysis population included 51 participants with 24 in the fluid-sparing and 27 in the usual care group. Consent for ongoing participation was obtained for 48/51 (94%) participants. Baseline characteristics of study participants are presented in **Table 1**.

**Figure 2:**
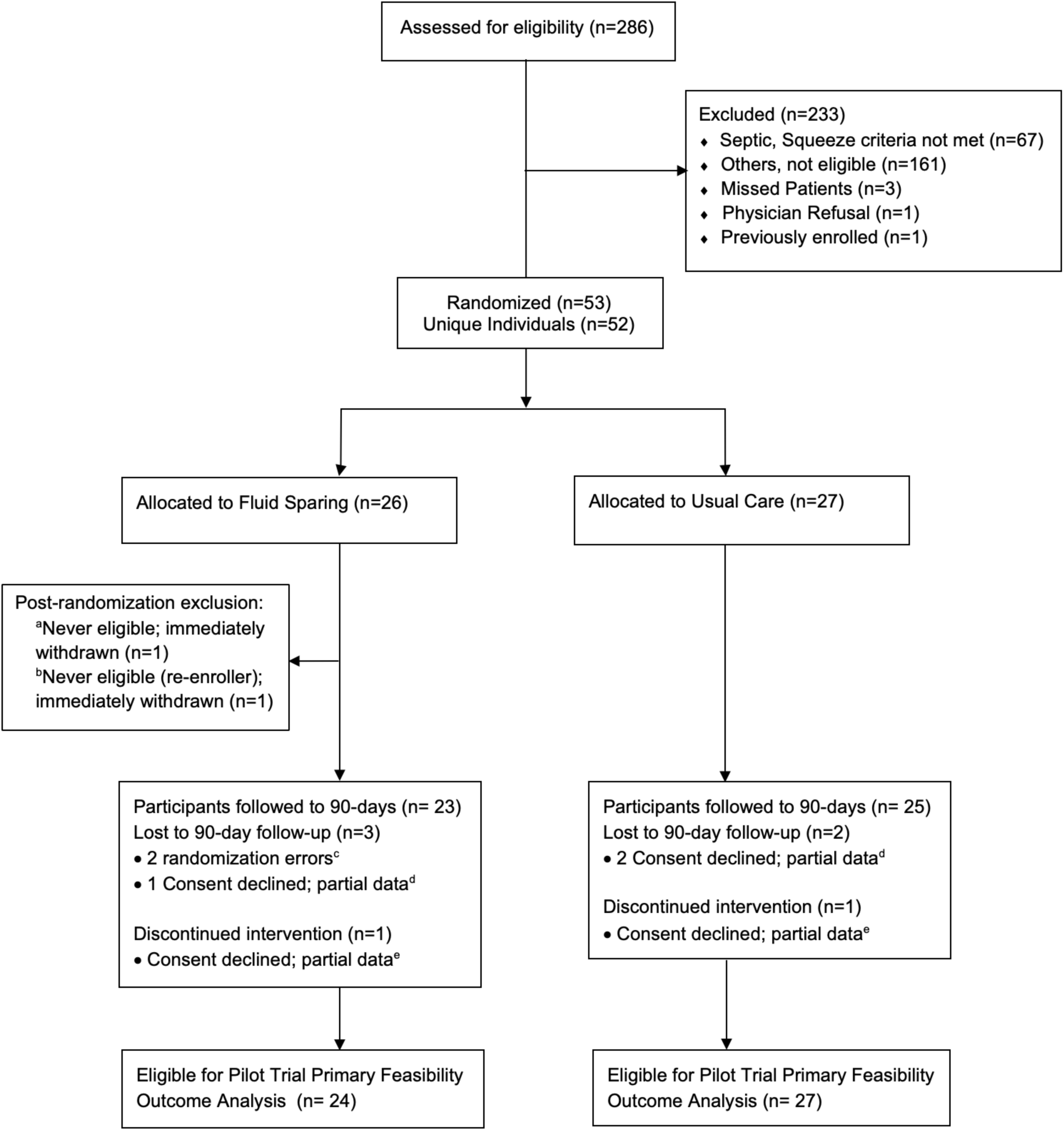
SQUEEZE Pilot Trial CONSORT Flow Diagram. **Legend for Figure 2.** a. This was an early randomization error shortly after recruitment began. An eager research team member misapplied the screening criteria and randomized a stable ward patient. Immediately recognized; the patient did not receive the intervention and was excluded. No other data is available. b. This randomization error occurred when a research team member failed to check the log of previously enrolled patients. The error was quickly recognized, the patient did not receive the intervention, and was excluded. The first enrolment was retained. c. The details of these randomization errors^a,b^ are noted above. d. Due to the exception to consent alternative consent model, participants enrolled may decline consent post-randomization. In such instances, data is retained up until the point of consent decline as described in our protocol. e. The intervention was only discontinued if it was still being applied at the time when consent was declined.

**Table 1.**
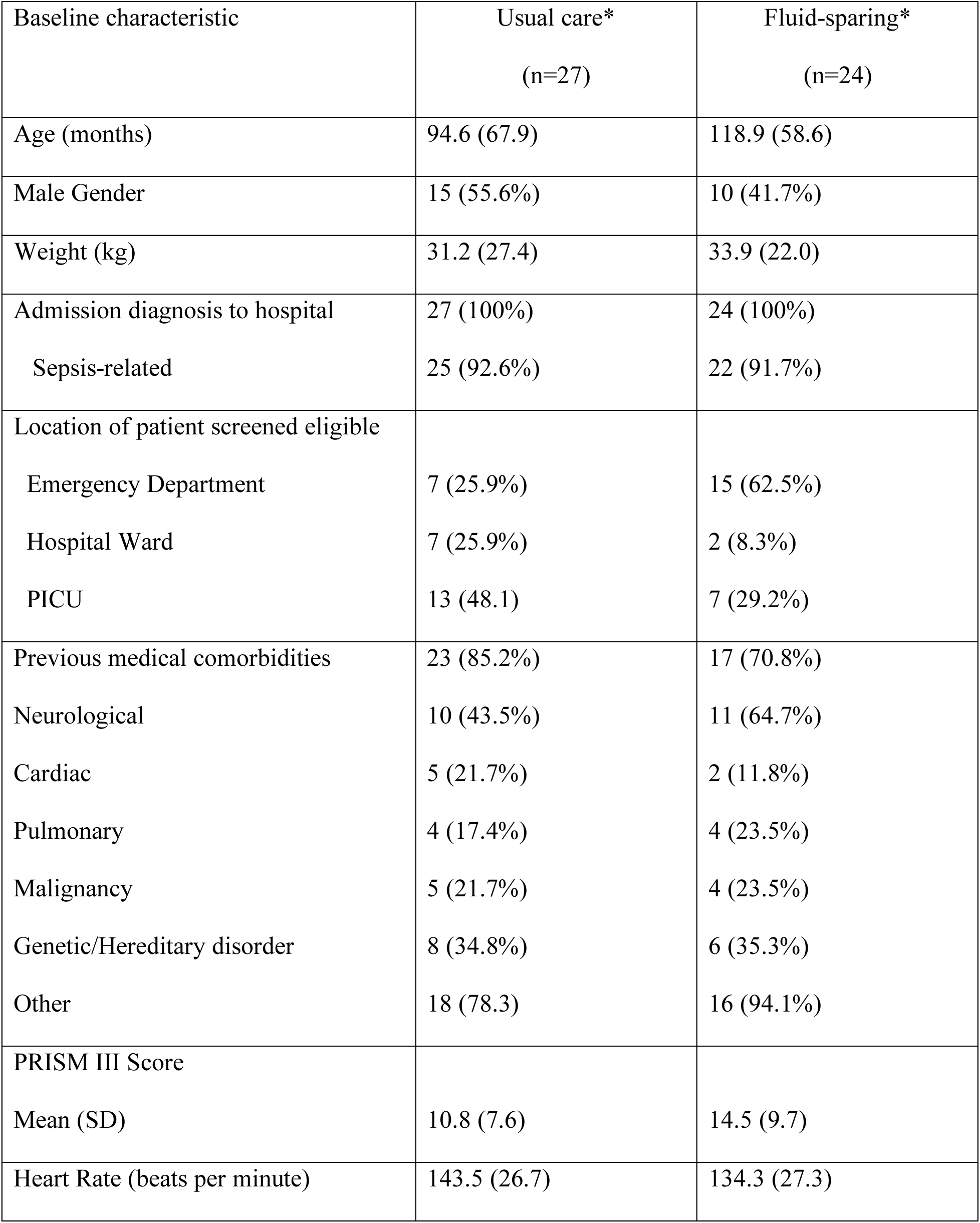
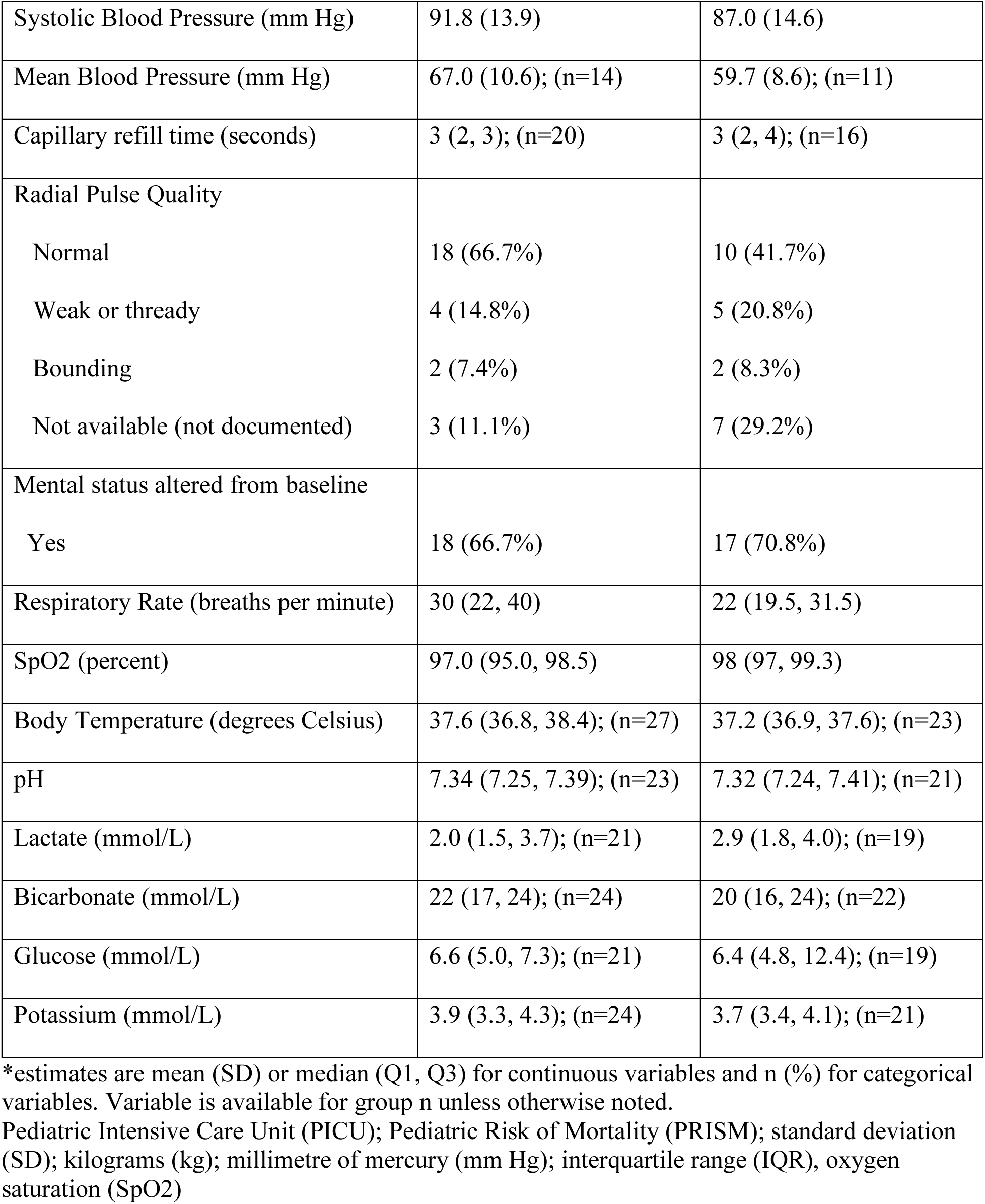
Baseline Characteristics.

SQUEEZE pilot trial feasibility outcomes are presented in **Table 2**. The enrolment rate was estimated at 1.8 (51/29); 95% C.I. 1.3-2.2 participants/month, which met our feasibility criteria. Importantly, we implemented an early amendment to one of the inclusion criteria based on our experience during the first 6 months of recruitment. We identified that our initial time window of 2 hours for the minimum fluid administration criteria to be met was too short and either delayed participant entry into the trial or resulted in exclusion of patients we believed should be included. A protocol amendment increasing this timeframe from 2 to 6 hours was implemented following REB approval in September of 2014 and appeared to impact the slope of recruitment for the remainder of the trial (**Figure 3**). The post amendment recruitment rate was estimated at 2.0 (41/21); 95% C.I. 1.4-2.6 participants/month.

**Figure 3.**
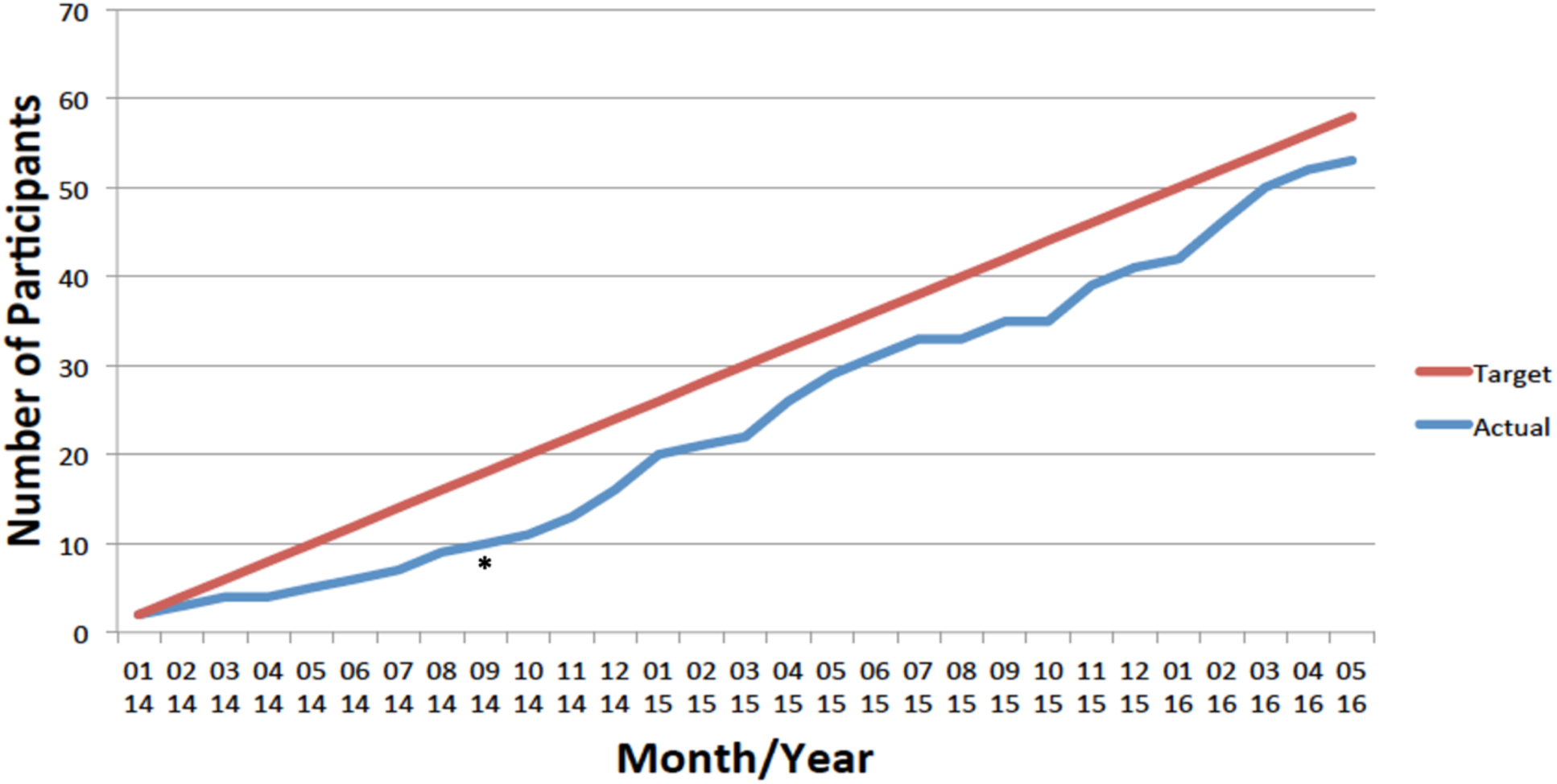
SQUEEZE Pilot Trial Recruitment **Legend for Figure 3.** SQUEEZE Pilot Trial Recruitment On the y-axis of the figure is the cumulative number of participants randomized into the trial. On the x-axis are the month and year of recruitment. Asterisk (*) denotes timing of implementation of a protocol amendment to an inclusion criteria in September, 2014. The amendment lengthened the time window from 2 to 6 hours within which the minimum volume of fluid bolus therapy required to meet eligibility criteria was received.

**Table 2.**
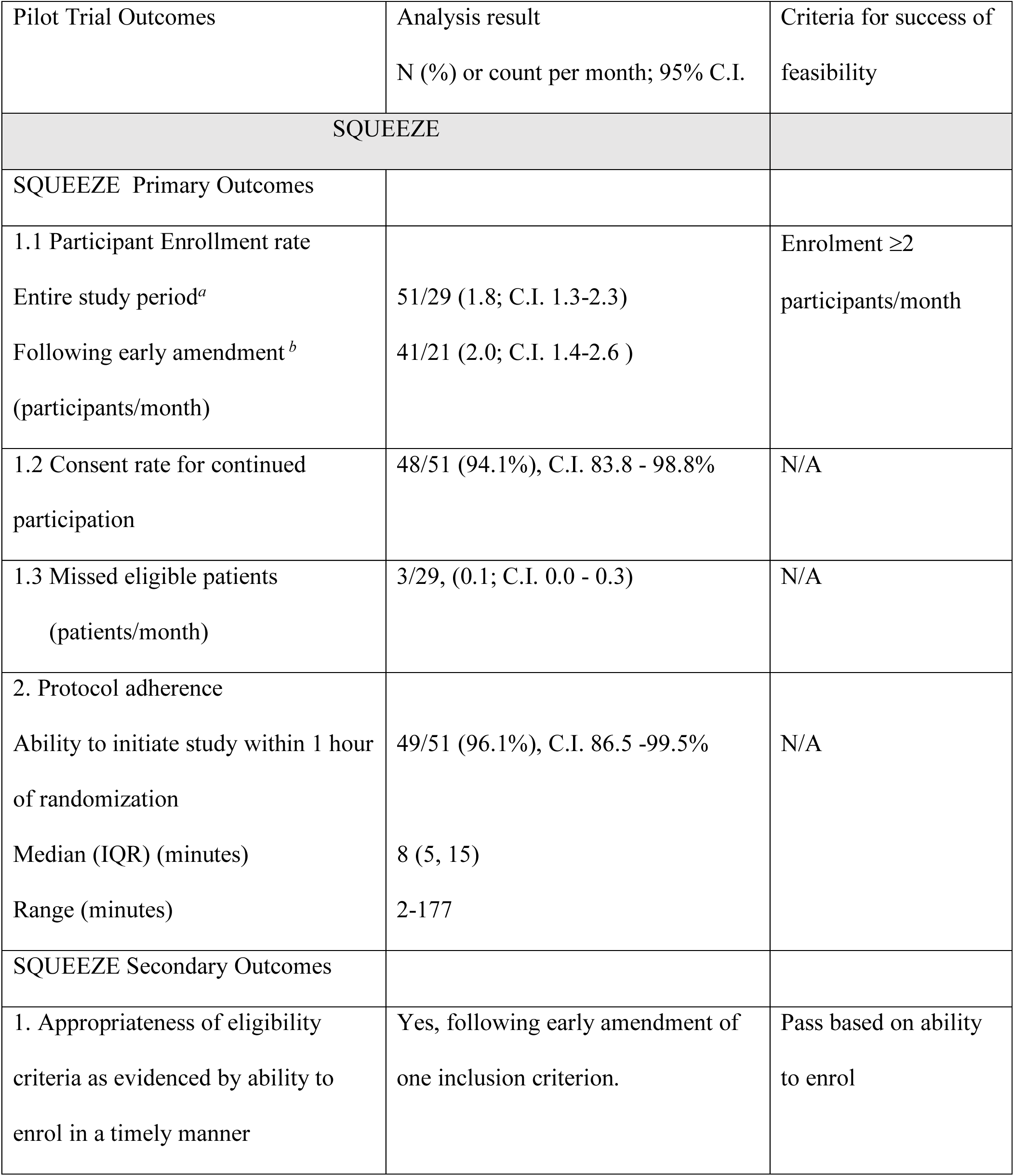
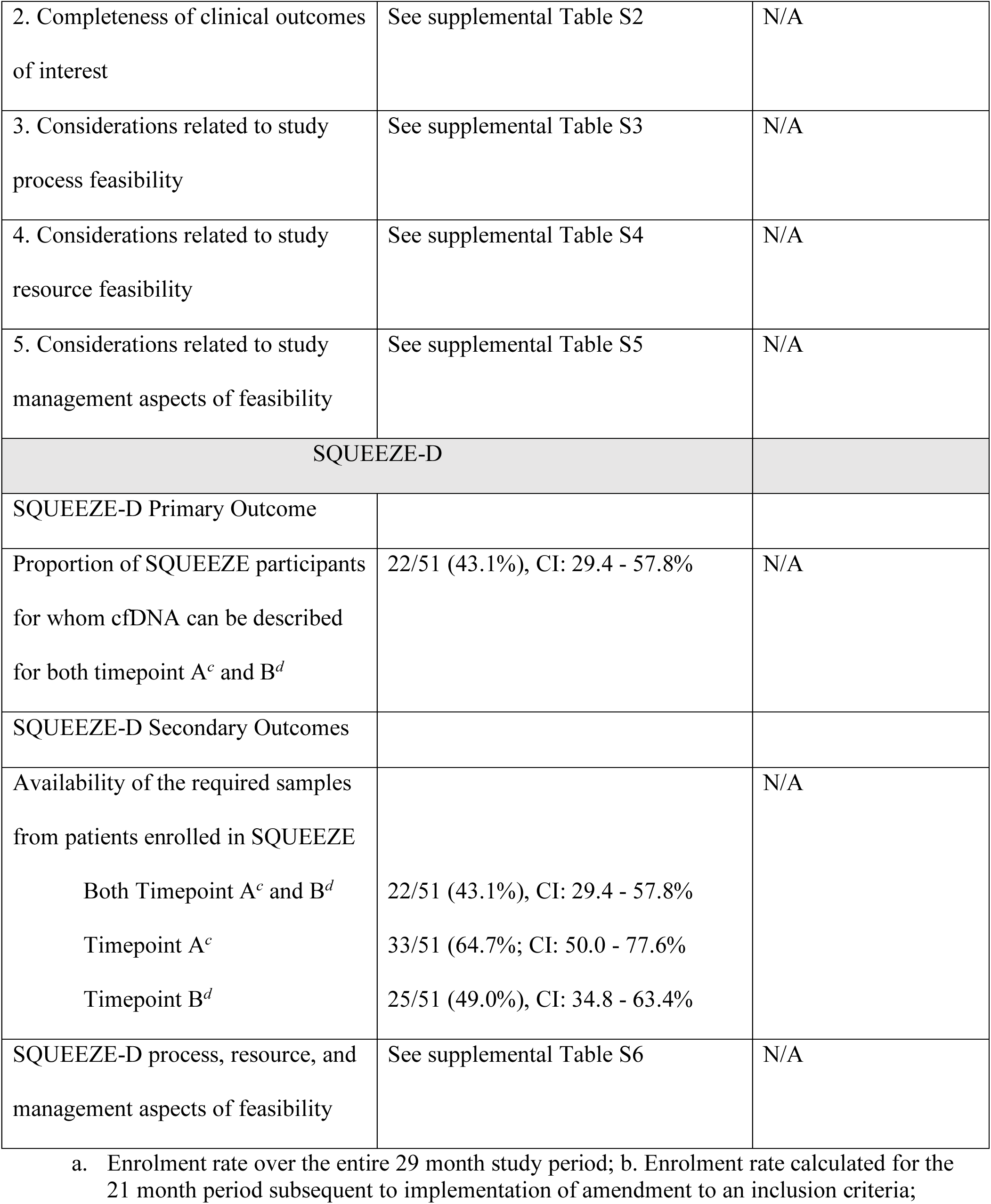
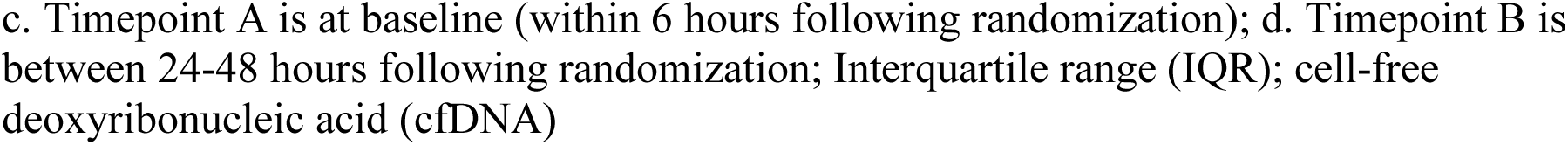
SQUEEZE pilot trial feasibility outcomes.

The median (IQR) time to initiate study procedures was 8 (5, 15) minutes, and implemented within 1 hour of randomization in 49/51 (96.1%; 95% C.I. 86.5 - 99.5%). In the two instances where this was not achieved, one involved a delay in reaching a member of the healthcare team while in the other study package implementation was delayed. The completeness of hemodynamic and clinical outcome data of interest for the full trial are presented in **Table S2**. Additional supplementary tables present secondary outcome findings related to study process (**Table S3**), resource (**Table S4**), and management (**Table S5**) aspects of feasibility and for SQUEEZE-D (**Table S6**). Our ability to collect pre-randomization (**Table S7**) and post-randomization (**Table S8**) data describing fluid and blood product intake and output, and hemodynamic descriptive data (**Table S9**) are also provided. Completeness of pre- and post-randomization descriptive data for culture results, antimicrobial therapy and laboratory data are presented in **Table S10** and **Table S11**.

For scientific aspects of feasibility, there were no adverse events or SAEs. Protocol deviations, violations, suspensions and withdrawals (reported in **Table S3)** are relevant from a scientific perspective due to their potential to impact the effectiveness of the intervention and to introduce risk of bias in the case of withdrawals. We assessed the effectiveness of the intervention to spare fluid at a single timepoint given the open label design of the trial. These results as presented in our 2016 application to CIHR for large multicentre trial funding are presented in **Figure 4**. It was determined that 400 participants (200 per arm) were required for a multicenter trial to detect an estimated 30% difference in the time to shock reversal based on a two-sided t-test of the null hypothesis that there is no difference between groups (based on geometric mean), with type 1 error (α) at 0.05 and power (β) at 80%. We considered this difference minimally clinically meaningful as this corresponded to approximately one nursing shift. It is important to note the use of the geometric mean for statistical testing. Use of geometric means for between group comparisons is appropriate when data may be skewed - a situation where use of group means would otherwise be inappropriate. An alternative approach in the setting of skewed outcome data could be comparison of group medians. While survival analysis was considered for the main trial, the sample size required for this analysis approach was prohibitive.

**Figure 4.**
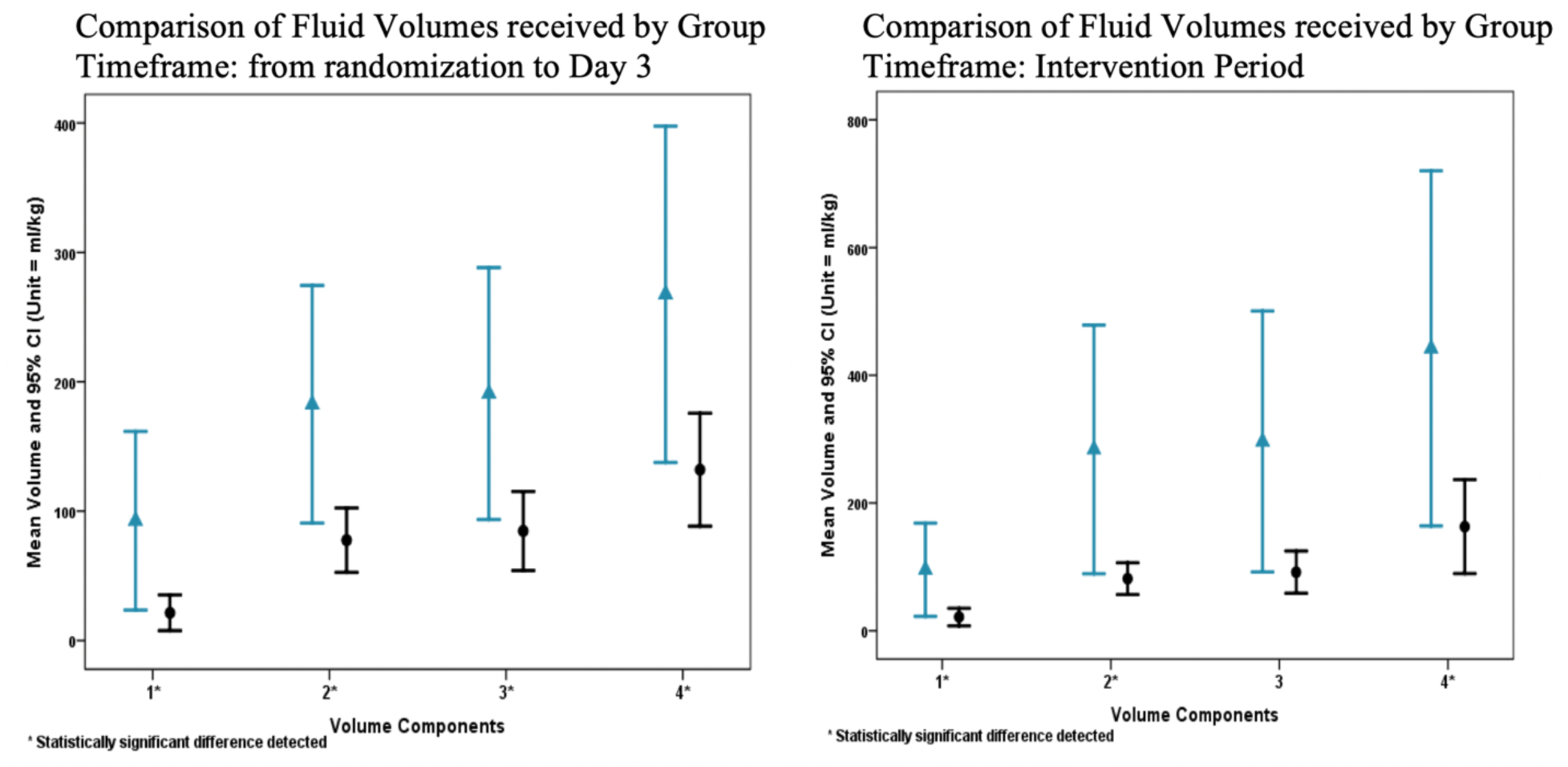
Between group separation in fluid volume received reported by volume components **Legend Figure 4.** 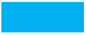 Group A (Usual Care) 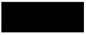Group B (Fluid-sparing) **Caption for Figure 4.** On the y-axis of the two figures - mean fluid volume received (mL/kg). On the x-axis of the two figures - fluid volume components: 1 - isotonic fluid boluses only, 2 - isotonic fluid boluses + maintenance fluid, 3 - isotonic fluid boluses + maintenance fluid + blood products, 4 - all fluid intake. Asterisk (*) - statistically significant between group difference at p<0.05 level. In the figure on the left, the timeframe is defined as the first six 12-hour periods (based on nursing shifts) following randomization, with the first shift a partial shift. In the figure on the right, the intervention period is defined as the period from randomization until shock is reversed for each participant.

## Discussion

We completed a single center pilot randomized controlled trial evaluating a fluid-sparing intervention vs usual care in children with a clinical diagnosis of septic shock and a need for ongoing resuscitation. The main finding of our study was that the protocol was feasible to conduct and this supported progression to a larger, multicentre trial based on our ability to enroll participants and rapidly implement study procedures. Our second main finding was that adherence to the protocol was effective in achieving between group separation in the volume of fluid received. Thirdly, our experience with study procedures and data collection informed refinement of the protocol and data collection plan for the main trial. Finally, our approach to operationalizing the exception to consent process used in the trial worked well and was generally well accepted.

In designing the SQUEEZE protocol, our aim was for the study to be pragmatic so that our findings would be widely generalizable. We therefore recruited from various locations within the hospital including the PED, inpatient wards, and PICU. We decided not to recruit from community hospital sites due to the rarity of pediatric septic shock in conjunction with the expense and challenges of training and maintaining the engagement of community physicians. Inwald et al attempted recruitment of a similar population from community hospitals during this timeframe and deemed their protocol infeasible.^24^ We decided against recruitment in the prehospital environment for similar reasons. Patients critically ill enough to require transfer to tertiary care retained the opportunity to be enrolled if they met eligibility criteria upon arrival. This was an important feature of our protocol which supported enrolment into either tier 1 or tier 2 of the intervention. We considered this analogous to the real world setting where clinicians implement a guideline according to a patient’s clinical status when they assume care.

Another important aspect of our eligibility criteria was the minimum required volume of fluid bolus therapy and the timeframe within which this was received. We selected a minimum volume of 40 mL/kg (2 litres for >50 kg) which corresponded to 2 fluid boluses because we believed this was necessary to select children at risk for relatively rare outcomes of interest. In comparison with two other pilot trials investigating interventions to spare fluid, we succeeded in enrolling sicker children as evidenced by participant PRISM III scores.^24,25^

Adherence to the SQUEEZE protocol was effective in achieving a significant fluid-sparing effect as shown in **Figure 4**. This was sustained to at least 72 hours, which we attribute to our intervention being applied until shock reversal was confirmed. In designing the SQUEEZE intervention, we believed that early initiation of vasoactive medication(s) while important, was insufficient to result in a meaningful difference in fluid volume received over an extended period of time. Prioritized use of vasoactive medications provided an alternative strategy to target recommended hemodynamic endpoints while avoiding or otherwise limiting further fluid bolus therapy. Protocol deviations occurred in a minority of participants and did not compromise between group separation in the volume of fluid received.

Testing our protocol and data collection plan provided significant learning opportunities. One unanticipated finding was that the telephone accessible randomization system required 24/7 was vulnerable to power outages. We therefore shifted to REDCap based randomization for the main trial. We also learned that having the study pager linked to the MET paging waterfall resulted in an excessive number of calls and high screening workload with the majority of these patients ineligible and so this was abandoned. USCOM assessments could not be reliably completed and were eliminated. Finally, our experience allowed us to refine our data collection plan for the main phase of the trial. Radial pulse quality was eliminated as this was not documented by nursing staff. We shifted from PRISM III to PRISM IV due to its improved reliability.^26^ Acute kidney injury was added as an outcome, as its exclusion from the pilot trial CRF was an oversight.

The SQUEEZE pilot trial was the first study in Canada to enroll children into a RCT using an exception to consent process. This alternative consent model provides a temporary waiver of consent to enable research in individual medical emergencies and allows enrolment, initiation of study procedures and data collection until such time as full informed consent (and assent as applicable) can be completed. We implemented this using a practical approach of providing a one page notice of enrolment to SDMs as early as possible to inform them of their child’s enrolment. This was followed by at minimum daily research team contact with the participant’s healthcare team to determine the most appropriate time to approach the SDMs for full informed consent discussions. Our approach was generally well accepted and this has informed other trials since SQUEEZE began.^27^

Our pilot trial had several limitations. We expected some post-randomization exit due to consent decline and that some outcomes for these participants may be unavailable which could compromise study validity.^28^ We did not anticipate randomization errors, however these are well described and in retrospect not surprising in a fast-paced resuscitation trial.^29^ Learnings from our experience with the pilot trial protocol were useful to inform site training for the large multicenter phase of the trial. Overall, our loss to follow-up rate was low. The very low number of missed patients reflects the high degree of interest in the trial and few gaps in research team on call coverage, which we expected may not be replicable at external sites. We did attempt to add a second site to evaluate external feasibility however the pilot trial recruitment target was reached before the external site could be added. Finally, our pilot trial took over 2 years to complete, reflecting the challenges of conducting interventional research in critically ill children.

## Conclusions

We determined that the multicenter SQUEEZE trial was feasible to conduct. Pilot trial data were essential to justify funding and inform conduct of the full-scale multicentre trial which is now complete. We believe the important lessons learned from this pilot trial will be of interest to readers given the complexities of conducting time-sensitive pediatric resuscitation research.

## Supporting information

SQUEEZE Pilot Trial Supplemental Digital File

## Data Availability

All data produced in the present study are available upon reasonable request to the authors.

## Acknowledgements

We would like to acknowledge methodological input into the SQUEEZE pilot trial protocol by Pediatric Emergency Research Canada and the Canadian Critical Care Trials Group, as well as ongoing input and advice during conduct of this trial. This manuscript also benefitted from critical review by the CCCTG Grants and Manuscript committee. We would like to thank the staff and trainees of the McMaster Children’s Hospital PED and PICU who embraced this pilot trial at a time (2014-2016) when use of exception to consent and investigation of a fluid-sparing intervention in children with septic shock were novel and as of yet untested.

## References

1. Brierley J, Carcillo JA, Choong K, et al. Clinical practice parameters for hemodynamic support of pediatric and neonatal septic shock: 2007 update from the American College of Critical Care Medicine. Critical Care Medicine 2009;37(2):666–88, doi:10.1097/CCM.0b013e31819323c6

2. Evans L, Rhodes A, Alhazzani W, et al. Surviving Sepsis Campaign: International Guidelines for Management of Sepsis and Septic Shock 2021. Crit Care Med 2021;49(11):e1063–e1143, doi:10.1097/CCM.0000000000005337

3. Maitland K, Kiguli S, Opoka RO, et al. Mortality after fluid bolus in African children with severe infection. New England Journal of Medicine 2011;364(26):2483–95

4. Boyd JH, Forbes J, Nakada TA, et al. Fluid resuscitation in septic shock: a positive fluid balance and elevated central venous pressure are associated with increased mortality. Critical Care Medicine 2011;39(2):259–65, doi:10.1097/CCM.0b013e3181feeb15

5. Vincent J-L, Sakr Y, Sprung CL, et al. Sepsis in European intensive care units: Results of the SOAP study*. Critical Care Medicine 2006;34(2):344–353, doi:10.1097/01.ccm.0000194725.48928.3a

6. Arikan AA, Zappitelli M, Goldstein SL, et al. Fluid overload is associated with impaired oxygenation and morbidity in critically ill children. Pediatric Critical Care Medicine 2012;13(3):253–8

7. Ford N, Hargreaves S, Shanks L. Mortality after fluid bolus in children with shock due to sepsis or severe infection: a systematic review and meta-analysis. PloS one 2012;7(8):e43953, doi:10.1371/journal.pone.0043953

8. Dwivedi DJ, Toltl LJ, Swystun LL, et al. Prognostic utility and characterization of cell-free DNA in patients with severe sepsis. Crit Care 2012;16(4):R151, doi:10.1186/cc11466

9. Liaw PC, Fox-Robichaud AE, Liaw KL, et al. Mortality Risk Profiles for Sepsis: A Novel Longitudinal and Multivariable Approach. Crit Care Explor 2019;1(8):e0032, doi:10.1097/CCE.0000000000000032

10. Consent for Research in Individual Medical Emergencies. Chapter 3, Section B, Article 3.8. In: Tri-Council Policy Statement: Ethical Conduct for Research Involving Humans. Canadian Institutes of Health Research, Natural Sciences and Engineering Research Council of Canada, Social Sciences and Humanities Research Council of Canada: Ottawa, Canada; 2014.

11. Chan AW, Tetzlaff JM, Altman DG, et al. SPIRIT 2013: new guidance for content of clinical trial protocols. Lancet 2013;381(9861):91–2, doi:10.1016/S0140-6736(12)62160-6

12. Chan AW, Tetzlaff JM, Gotzsche PC, et al. SPIRIT 2013 explanation and elaboration: guidance for protocols of clinical trials. BMJ 2013;346(e7586, doi:10.1136/bmj.e7586

13. Parker MJ, Thabane L, Fox-Robichaud A, et al. A trial to determine whether septic shock-reversal is quicker in pediatric patients randomized to an early goal-directed fluid-sparing strategy versus usual care (SQUEEZE): study protocol for a pilot randomized controlled trial. Trials 2016;17(1):556, doi:10.1186/s13063-016-1689-2

14. Eldridge SM, Chan CL, Campbell MJ, et al. CONSORT 2010 statement: extension to randomised pilot and feasibility trials. Pilot and Feasibility Studies 2016;2(1), doi:10.1186/s40814-016-0105-8

15. ARISE Investigators, ANZICS Clinical Trials Group, Peake SL, Delaney A, Bailey M, Bellomo R, et al. Goal-directed resuscitation for patients with early septic shock. N Engl J Med 2014;371(16):1496–506, doi:10.1056/NEJMoa1404380

16. Thabane L, Ma J, Chu R, et al. A tutorial on pilot studies: the what, why and how. BioMed Central Medical Research Methodology 2010;10(1), doi:10.1186/1471-2288-10-1

17. Harris PA, Taylor R, Thielke R, et al. Research electronic data capture (REDCap)--a metadata-driven methodology and workflow process for providing translational research informatics support. J Biomed Inform 2009;42(2):377–81, doi:10.1016/j.jbi.2008.08.010

18. Cocks K, Torgerson D. Sample size calculations for pilot randomized controlled trials: a confidence interval approach. Journal of Clinical Epidemiology 2013;66(197-201

19. Cook D, Lauzier F, Rocha MG, et al. Serious adverse events in academic critical care research. CMAJ 2008;178(9):1181–1184

20. Hamilton Integrated Research Ethics Board. Instructions for Reporting of Local and Non-Local Adverse Events, Version: November 2015_B. (2015). Available at: https://hireb.ca/wp-content/uploads/2023/03/Serious-Adverse-Events-SAE-Unanticipated-Problems.pdf (Accessed: April 5, 2024).

21. Guidance for Clinical Trial Sponsors: Establishment and Operation of Clinical Trial Data Monitoring Committees. OMB Control No. 0910-0581. U.S. Department of Health and Human Services Food and Drug Administration. 2006.

22. Shulz KF, Altman DG, Moher D, et al. Consort 2010 Statement: Updated guidelines for reporting parallel group randomized trials. Journal of clinical epidemiology 2010;63(8):834–40, doi:10.1016/j.jclinepi.2010.02.005

23. Moher D, Hopewell S, Shulz KF, et al. Consort 2010 Explanation and Elaboration: Updated guidelines for reporting parallel group randomised trials. Journal of clinical epidemiology 2010;63(8):e1-e37, doi:10.1016/j.jclinepi.2010.03.004

24. Inwald DP, Canter R, Woolfall K, et al. Restricted fluid bolus volume in early septic shock: results of the Fluids in Shock pilot trial. Arch Dis Child 2019;104(5):426–431, doi:10.1136/archdischild-2018-314924

25. Harley A, George S, Phillips N, et al. Resuscitation With Early Adrenaline Infusion for Children With Septic Shock: A Randomized Pilot Trial. Pediatr Crit Care Med 2024;25(2):106–117, doi:10.1097/PCC.0000000000003351

26. Pollack MM, Holubkov R, Funai T, et al. The Pediatric Risk of Mortality Score: Update 2015. Pediatr Crit Care Med 2016;17(1):2–9, doi:10.1097/PCC.0000000000000558

27. Parker MJ, de Laat S, Schwartz L. Exploring the experiences of substitute decision-makers with an exception to consent in a paediatric resuscitation randomised controlled trial: study protocol for a qualitative research study. BMJ Open 2016;6(9):e012931, doi:10.1136/bmjopen-2016-012931

28. Akl EA, Briel M, You JJ, et al. Potential impact on estimated treatment effects of information lost to follow-up in randomised controlled trials (LOST-IT): systematic review. BMJ 2012;344(e2809, doi:10.1136/bmj.e2809

29. Yelland LN, Sullivan TR, Voysey M, et al. Applying the intention-to-treat principle in practice: Guidance on handling randomisation errors. Clin Trials 2015;12(4):418–23, doi:10.1177/1740774515588097

30. Parker M. SQUEEZE fluid-sparing (intervention) and usual care (control) management strategies. MacSphere; 2024. Available at: http://hdl.handle.net/11375/29921 (Accessed: June 29, 2024)

