## Supplementary material for "The SQUEEZE pilot trial: a trial to determine whether septic shock reversal is quicker in pediatric patients randomized to an early goal directed fluid-sparing strategy vs. usual care": SQUEEZE Pilot Trial Supplemental Digital File

### SQUEEZE Pilot Trial Supplemental Digital Content

ClinicalTrials.gov [NCT01973907]

Parker MJ, Choong K, Fox-Robichaud A, Liaw PC, Thabane L, for the Canadian Critical Care Trials Group and the Canadian Critical Care Translational Biology Group.

Table of Contents


| | | |
|---|---|---|
| Table S1 | Summary of Protocol Versions and Amendments | pg 3 |
| Table S2 | Hemodynamic, Clinical and Adverse Event Outcomes | pg 4 |
| Table S3 | Study process feasibility outcomes | pg 5 |
| Table S4 | Study resource feasibility outcomes | pg 6 |
| Table S5 | Study management feasibility outcomes | pg 7 |
| Table S6 | SQUEEZE-D secondary feasibility outcomes | pg 8 |
| Table S7 | Completeness of pre-randomization fluid and blood product data | pg 9 |
| Table S8 | Completeness of post-randomization fluid and blood product data | pg 10 |
| Table S9 | Completeness of hemodynamic descriptive data | pg 11 |
| Table S10 | Completeness of pre-randomization culture and antimicrobial data | pg 12 |
| Table S11 | Completeness of post-randomization culture, antimicrobial and laboratory data | pg 13 |
| Appendix S1 | CONSORT checklist for pilot and feasibility trials | pg 14 |
| Appendix S2 | Table 1 - Extended. Baseline Characteristics | pg 17 |

Table S1. Summary of Protocol Versions and Amendments

| Protocol Version/Date | Date REB Approved | Key Details |
|---|---|---|
| Version 1; 04/04/2013 | n/a | Provisional approval - revisions requested by REB |
| Version 2;20/05/2013 | 04/06/2013 | Revised protocol received full approval |
| Version 3; 25/06/2013 | 03/07/2013 | Edits to protocol prior to trial launch |
| Version 4; 27/10/2013 | 18/11/2013 | Edits to protocol prior to trial launch |
| **RECRUITMENT OPENS Jan 6, 2014** | | |
| **First Participant Enrolled Jan 7, 2014** | | |
| Version 5; 09/07/2014 | 19/08/2014 | Lengthening of time window from 2 to 6 hours within which minimum volume of isotonic fluid bolus therapy can be received (inclusion criteria)<br>Additional Translational study (PERSEVERE) added |
| Version 6; 25/11/2014 | 16/12/2014 | Approval to add second external site to pilot trial<br>Funding sources (new) added |

Table S2: Completeness of Hemodynamic and Clinical Outcome and Adverse Event Data

| Outcome | Able to collect, N (%), 95% C.I. (N=51) |
|---|---|
| **Hemodynamic Outcome** | |
| Time to Shock reversal (hours) | 49 (96.1%), C.I. 86.5 -99.5% |
| **Complications possibly attributable to fluid overload or third spacing** | |
| Pleural effusion requiring drainage (Y/N) | 50 (98.0%), C.I. 89.6 - 100% |
| Intra-abdominal hypertension (Y/N) | 50 (98.0%), C.I. 89.6 - 100% |
| Highest bladder pressure (mm Hg) | 50 (98.0%), C.I. 89.6 -100% |
| Abdominal compartment syndrome (Y/N) | 49 (96.1%), C.I. 86.5 - 99.5% |
| Soft tissue edema (rank: none, mild, moderate, severe) | 51 (100%), C.I. 93.0 - 100% |
| Pulmonary edema (Y/N) | 50 (98.0%), C.I. 89.6 - 100% |
| Lasix (furosemide) exposure (Y/N) | 50 (98.0%), C.I. 89.6 - 100% |
| Lasix (furosemide) exposure (mg/kg) | 50 (98.0%), C.I. 89.6 - 100% |
| Other diuretics used (Y/N) | 50 (98.0%), C.I. 89.6 - 100% |
| **Complications possibly attributable to inotrope/vasopressor use** | |
| Clinical signs of digital or soft tissue ischemia (Y/N) | 50 (94.1%), C.I. 89.6 - 100% |
| Digital ischemia requiring revision amputation (Y/N) | 48 (94.1%), 95% C.I. 83.8 - 98.8% |
| Clinical signs of compromised bowel perfusion (Y/N) | 48 (94.1%), 95% C.I. 83.8 - 98.8% |
| **Clinical Course and Procedure Outcomes** | |
| Length of Hospital Stay (days) | 48 (94.1%), 95% C.I. 83.8 - 98.8% |
| PICU admission (Y/N) | 51 (100%), C.I. 93.0 - 100% |
| Length of PICU stay (days) | 48 (94.1%), 95% C.I. 83.8 - 98.8% |
| PELOD score components (Y/N) | 49 (96.1%), 95% C.I. 86.5 -99.5% |
| PELOD-2 score components (Y/N) | 49 (96.1%), 95% C.I. 86.5 -99.5% |
| Data to calculate of Ventilator Free Days (Y/N) | 48 (94.1%), 95% C.I. 83.8 - 98.8% |
| 28-day Mortality (Y/N) | 48 (94.1%), 95% C.I. 83.8 - 98.8% |
| Hospital Mortality (Y/N) | 48 (94.1%), 95% C.I. 83.8 - 98.8% |
| Cause of Death (if applicable) | 48 (94.1%), 95% C.I. 83.8 - 98.8% |
| Arterial Line Placement (Y/N) | 51 (100%), C.I. 93.0 - 100% |
| Central Line placement (Y/N) | 51 (100%), C.I. 93.0 - 100% |
| Renal Replacement Therapy (Y/N) | 49 (96.1%), 95% C.I. 86.5 -99.5% |
| Chest tube (Y/N) | 50 (98.0%), C.I. 89.6 -100% |
| Peritoneal Drain (Y/N) | 49 (96.1%), 95% C.I. 86.5 -99.5% |
| Echocardiography performed (Y/N) | 51 (100%), C.I. 93.0 - 100% |
| Patient placed on mechanical circulatory support, such as ECMO, to treat refractory shock (Y/N) | 49 (96.1%), 95% C.I. 86.5 -99.5% |

PELOD - Pediatric Logistic Organ Dysfunction, ECMO - extracorporeal membrane oxygenation, PICU - Pediatric Intensive Care Unit

Table S3. Considerations related to study process feasibility

| Process Feasibility Item | Descriptive finding (N=51) |
|---|---|
| Time from earliest eligibility to randomization | |
| Median (IQR) (minutes) | 23 (9.5, 46) |
| Range (minutes) | 0-1080 |
| Time from randomization to communication of allocation | |
| Median (IQR) (minutes) | 5 (2.5, 10) |
| Range (minutes) | 1-58 |
| Healthcare team member receiving allocation communication | |
| Staff physician | 29 (56.9%) |
| Registered Nurse | 1 (2.0%) |
| PICU fellow | 20 (39.2%) |
| PICU resident | 1 (2.0%) |
| Time from communication of allocation to initiation of study procedures | |
| Median (IQR) (minutes) | 2 (1, 5) |
| Range (minutes) | 0 - 175 |
| Time from randomization to initiation of study procedures | |
| Median (IQR) (minutes) | 8 (5, 15) |
| Range (minutes) | 2 - 177 |
| Protocol acceptability to medical staff | |
| Tier 1, N (%) | 51 (100%) |
| Tier 2, N (%) | 51 (100%) |
| Protocol Deviations | |
| Study procedures not implemented within 1 hr of randomization | 2/51 (3.9%) |
| Any fluid bolus >10 mL/kg (500 mL if >50 kg) administered to participant assigned to fluid-sparing arm | 6/24 (25%) |
| Any fluid bolus of 20 mL/kg (1000 mL if > 50 kg) administered to participant assigned to fluid-sparing arm | 3/24 (12.5%) |
| Vasoactive medications not initiated in fluid sparing arm | 4/24 (16.7%) |
| Notification of enrolment sheet not provided | 2/51 (3.9%) |
| Notification not provided within 24 hours | 14/51 (27.5%) |
| Protocol violations (with reasons) | |
| PICU fellow was asked to approach family to discuss study after hours while child still unstable; research team not consulted | 1/51 (2.0%) |
| Protocol suspensions (with reasons) | |
| Protocol suspended while participant in the operating room | 6/51 (11.8%) |
| Protocol Withdrawals (with reasons) | |
| Consent for ongoing participation declined | 3/51 (5.9%) |
| Other | Telephone access to randomization system |

Table S4. Considerations related to study resource feasibility

| Resource feasibility item | Descriptive finding |
|---|---|
| Daily screening of PICU admitted patients and MET Team activations, including for missed patients | 1 hour/day |
| Realtime screening when study team contacted (per case) | 15-30 minutes/case |
| Randomization, notification of clinical team, implementation of study procedures | 1 hour/case |
| Healthcare staff cooperation with study protocol | |
| Yes | 50/51 (98.0%) |
| No | 1/51 (2.0%) |
| Ability to perform USCOM assessments as specified per protocol | 0 |
| USCOM Equipment failure rate | 0 |
| Time to complete consent process | Variable; multiple interactions often required 1.5 hrs/case on average |
| Time to complete case report form during intervention period | 3 hours/case; 2-4 hours typical |
| Follow-up data collection to 90 days | 2 hours/case |
| Time to complete REDCap data entry from paper CRF | 2 hours/case |
| Study promotion activities: Attend PICU daily Mon-Fri; Visit Emergency Department 3x/week | 1.5 hours/week |

Table S5. Considerations related to study management aspects of feasibility

| Management feasibility item | Descriptive finding |
| --- | --- |
| On-call schedule to support 24/7 enrolment | Payment for on-call coverage required<br>Site investigators occasionally needed to provide call coverage |
| Use of pager to contact research team | Worked well, high reliability |
| Pager linked to MET team paging waterfall | Resulted in an excessive number of pages for non-sepsis MET team activations |
| Personnel availability for USCOM assessment | Personnel frequently unavailable when assessment required per protocol |
| REDCap database | worked well for data entry |
| Rate of successful data abstraction | Some clinical data of interest not reliably documented e.g. radial pulse quality |
| Rate of successful data entry | No problems with data entry into REDCap database from paper CRF. |
| Research Ethics Board review timeline<br>Lead site REB approval<br><br>External site REB approval | <br>2 months from initial submission to final approval<br>Lengthy review process at a single external site; pilot trial completed recruitment to target before external site approval obtained |

Table S6. SQUEEZE-D process, resource, management aspects of feasibility

| Process Feasibility | Descriptive Finding |
|---|---|
| Access to study specimen tubes | CPT tubes included in same study package as documents for clinical trial implementation |
| Specimen collection | Coordinated with clinical bloodwork ordered<br>Sample collected by participant's nurse |
| Specimen transport for initial processing | Specimen transported to core laboratory with other clinically indicated bloodwork |
| Initial specimen processing | Core laboratory processed sample and placed in -80 C research freezer within Core lab |
| Specimen transport from clinical site to research laboratory | Samples shipped on dry ice to research lab at TaARI. Transferred to -80 C research freezer |
| Specimen storage | Initial storage: -80 C research freezer at McMaster Children's Hospital Core Lab<br>TaARI storage: -80 C research freezer in the laboratory of Drs. Fox-Robichaud and Liaw |
| Resource Feasibility | |
| Study specimen form completion | Research staff or investigator typically needed to complete in real-time 24/7; not feasible for main trial |
| Contemporaneous completion and faxing of SQUEEZE-D requisition to pair with clinical sample | Realtime fax and follow-up by phone-call to Core lab required to ensure clinical sample spun and frozen at -80 C within 1 hour. |
| Study process time | Not feasible to continue having study staff/investigators call Core Laboratory 24/7 |
| Availability of PICU clinical staff | Sample sometimes not collected when clinical staff (PICU nurses) busy with sick patient |
| Availability of Core Laboratory staff | Core laboratory staff busy, and staffing further decreased after hours. Prompting required to ensure sample processing. |
| Time-sensitive sample processing | Processing of fresh sample and placement in -80 freezer time sensitive for cfDNA (must be spun and frozen within 1 hour) |
| CPT tubes | Commonly available, inexpensive |
| CPT tube labels | Several specimens lost due to failure of label adherence in -80 freezer |
| Management feasibility | |
| Specimen form process | Current process not feasible to continue in main trial; new process required |

Table S7. Completeness of pre-randomization fluid and blood product data*

| Pre-randomization Fluid Data | Able to collect, N (%), 95% C.I. (N=51) |
|---|---|
| Fluid Bolus Therapy | |
| Date and time of first fluid bolus | 51 (100%), C.I. 93.0 - 100% |
| Normal Saline Fluid Bolus received (Y/N) | 51 (100%), C.I. 93.0 - 100% |
| Normal Saline boluses received (number of boluses) | 51 (100%), C.I. 93.0 - 100% |
| Total volume of Normal Saline as fluid bolus therapy (mL) | 51 (100%), C.I. 93.0 - 100% |
| Ringer's Lactate Fluid Bolus received (Y/N) | 51 (100%), C.I. 93.0 - 100% |
| Ringer's Lactate boluses received (number of boluses) | 51 (100%), C.I. 93.0 - 100% |
| Total volume of Ringer's Lactate as fluid bolus therapy (mL) | 51 (100%), C.I. 93.0 - 100% |
| 5% Albumin boluses received (Y/N) | 51 (100%), C.I. 93.0 - 100% |
| 5% Albumin boluses received (number of boluses) | 51 (100%), C.I. 93.0 - 100% |
| Total volume of 5% Albumin as fluid bolus therapy (mL) | 51 (100%), C.I. 93.0 - 100% |
| Blood Products | |
| Red Blood Cells received (Y/N) | 51 (100%), C.I. 93.0 - 100% |
| Total volume of red blood cells received (mL) | 51 (100%), C.I. 93.0 - 100% |
| Platelets received (Y/N) | 51 (100%), C.I. 93.0 - 100% |
| Total volume of platelets received (mL) | 51 (100%), C.I. 93.0 - 100% |
| 5% Albumin received as transfusion, not bolus (Y/N) | 51 (100%), C.I. 93.0 - 100% |
| Total volume of 5% Albumin as transfusion, not bolus (mL) | 51 (100%), C.I. 93.0 - 100% |
| 25% Albumin received (Y/N) | 51 (100%), C.I. 93.0 - 100% |
| Total volume of 25% albumin received (mL) | 51 (100%), C.I. 93.0 - 100% |
| Fresh Frozen Plasma received (Y/N) | 51 (100%), C.I. 93.0 - 100% |
| Total volume of Fresh Frozen Plasma received (mL) | 51 (100%), C.I. 93.0 - 100% |
| Cryoprecipitate received (Y/N) | 51 (100%), C.I. 93.0 - 100% |
| Total volume of Cryoprecipitate received (mL) | 51 (100%), C.I. 93.0 - 100% |

*Data collection for the period 24 hours prior to randomization

Table S8. Completeness of Fluid Intake, Fluid Output, and Fluid Bolus data post-randomization

| Fluid data to be collected | Able to collect, N (%), 95% C.I. (N=51) |
|---|---|
| Fluid Intake | |
| IV maintenance fluids (mL) | 49/51 (100%), C.I. 86.5 - 99.5% |
| IV total parenteral nutrition (mL) | 49/51 (100%), C.I. 86.5 - 99.5% |
| IV fluid bolus therapy (mL) | 49/51 (100%), C.I. 86.5 - 99.5% |
| Enteral fluid and nutrition (mL) | 49/51 (100%), C.I. 86.5 - 99.5% |
| Red Blood Cells (mL) | 49/51 (100%), C.I. 86.5 - 99.5% |
| Platelets (mL) | 49/51 (100%), C.I. 86.5 - 99.5% |
| 5% Albumin as transfusion, not bolus (mL) | 49/51 (100%), C.I. 86.5 - 99.5% |
| 25% Albumin (mL) | 49/51 (100%), C.I. 86.5 - 99.5% |
| Fresh frozen plasma (mL) | 49/51 (100%), C.I. 86.5 - 99.5% |
| Cryoprecipitate (mL) | 49/51 (100%), C.I. 86.5 - 99.5% |
| Intravenous Immunoglobulin (IVIG) (mL) | 49/51 (100%), C.I. 86.5 - 99.5% |
| IV medication administration (mL) | 49/51 (100%), C.I. 86.5 - 99.5% |
| Scheduled IV fluid replacement (mL) | 49/51 (100%), C.I. 86.5 - 99.5% |
| Fluids administered in the operating room (mL) | 49/51 (100%), C.I. 86.5 - 99.5% |
| Total fluid intake (mL) | 49/51 (100%), C.I. 86.5 - 99.5% |
| Fluid Losses or Removal | |
| Urine output (mL) | 49/51 (96.1%), C.I. 86.5 - 99.5% |
| Output from drains (mL) | 49/51 (96.1%), C.I. 86.5 - 99.5% |
| Renal replacement therapy (net fluid removal in mL) | 49/51 (96.1%), C.I. 86.5 - 99.5% |
| Losses as vomit/stool (mL) | 49/51 (96.1%), C.I. 86.5 - 99.5% |
| Total fluid output (mL) | 49/51 (96.1%), C.I. 86.5 -99.5% |
| Fluid Bolus therapy data for each fluid bolus event | |
| Fluid bolus administered (Y/N) | 49/51 (96.1%), C.I. 86.5 - 99.5% |
| Fluid bolus volume (mL) | 49/51 (96.1%), C.I. 86.5 - 99.5% |
| Fluid bolus volume (mL/kg) - using patient weight | 49/51 (96.1%), C.I. 86.5 - 99.5% |
| Fluid type (NS, RL, or 5% Albumin as bolus) | 49/51 (96.1%), C.I. 86.5 - 99.5% |
| Justification for fluid bolus form (fluid-sparing arm only) | 23/24 (95.8%), C.I. 78.9 - 99.9% |

IV - intravenous, NS - 0.9 Saline, RL - Ringer's Lactate

Table S9. Completeness of hemodynamic and related descriptive data

| Data to be collected | Able to collect , N (%), 95% C.I. (N=51) |
|---|---|
| **Hemodynamic and related data collected at baseline and every 12-hours** | |
| Highest heart rate (bpm) | 49 (100.0%), C.I. 86.5 -99.5% |
| Systolic blood pressure (mm Hg) | 49 (100.0%), C.I. 86.5 -99.5% |
| Diastolic blood pressure (mm Hg) | 49 (100.0%), C.I. 86.5 -99.5% |
| Mean blood pressure (mm Hg) | 25 (49.0%), C.I. 34.8 - 63.4% |
| Highest central venous pressure (mm Hg) | 49 (100%), C.I. 86.5 -99.5% |
| Lowest central venous pressure (mm Hg) | 49 (100%), C.I. 86.5 -99.5% |
| Central venous catheter location | 51 (100%), C.I. 93.0 - 100% |
| Invasive Mechanical ventilation (Y/N) | 51 (100%), C.I. 93.0 -100% |
| Highest mean airway pressure (mm Hg) | 49 (96.1%), C.I. 86.5 - 99.5% |
| **Data related to inotrope/vasopressor use** | |
| Vasoactive medications received (Y/N) | 51 (100%), C.I. 93.0 - 100% |
| First Inotrope/vasopressor infusion administered | 51 (100%), C.I. 93.0 - 100% |
| Inotrope/vasopressor dose data | 49 (96.1%), C.I. 86.5 - 99.5% |
| Dopamine (mcg/kg/min)<br>Dobutamine (mcg/kg/min)<br>Epinephrine (mcg/kg/min)<br>Norepinephrine (mc/kg/min)<br>Phenylephrine (mcg/kg/min)<br>Milrinone (mcg/kg/min) | 49 (96.1%), C.I. 86.5 -99.5%<br>49 (96.1%), C.I. 86.5 -99.5%<br>49 (96.1%), C.I. 86.5 -99.5%<br>49 (96.1%), C.I. 86.5 -99.5%<br>49 (96.1%), C.I. 86.5 -99.5%<br>49 (96.1%), C.I. 86.5 -99.5% |
| Data to support calculation of Vasoactive Inotropic Score | 49 (96.1%), C.I. 86.5 -99.5% |
| **USCOM data** | |
| USCOM study performed at Event-00 | 4 (7.8%), C.I. 2.2 - 18.9% |
| USCOM study performed at any timepoint | 14 (27.5%), C.I. 15.9 - 41.4% |
| USCOM study performed for ≥50% of required events | 1 (2%), C.I. 0.0 - 10.5% |
| **Other** | |
| Steroid administration (IV) | 51 (100%), C.I. 93.0 - 100% |

USCOM - Ultrasound Cardiac Output Monitor

Table S10. Completeness of pre-randomization culture and antimicrobial therapy data*

| Data to be collected | Able to collect, N (%), 95% C.I. (N=51) |
|---|---|
| Positive cultures | 51 (100%), C.I. 93.0 - 100% |
| Antimicrobials received | 51 (100%), C.I. 93.0 - 100% |

*culture and antimicrobial therapy data collected for the 24-hour period prior to randomization

Table S11. Completeness of post-randomization culture, antimicrobial, and laboratory data

| Data to be collected | Able to collect, N (%), 95% C.I. (N=51) |
|---|---|
| Positive cultures | 49 (96.1%), C.I. 86.5 -99.5% |
| Antimicrobial therapy | 49 (96.1%), C.I. 86.5 -99.5% |
| Laboratory data | 49 (96.1%), C.I. 86.5 -99.5% |

Appendix S1. CONSORT Checklist for Pilot and Feasibility Trials

#### CONSORT 2010 checklist of information to include when reporting a pilot or feasibility trial*

| Section/Topic | Item No | Checklist item | Reported on page No |
|---|---|---|---|
| **Title and abstract** | | | |
| | 1a | Identification as a pilot or feasibility randomised trial in the title | 1 |
| | 1b | Structured summary of pilot trial design, methods, results, and conclusions (for specific guidance see CONSORT abstract extension for pilot trials) | 2 |
| **Introduction** | | | |
| Background and objectives | 2a | Scientific background and explanation of rationale for future definitive trial, and reasons for randomised pilot trial | 3-4 |
| | 2b | Specific objectives or research questions for pilot trial | 3-4,7 |
| **Methods** | | | |
| Trial design | 3a | Description of pilot trial design (such as parallel, factorial) including allocation ratio | 4 |
| | 3b | Important changes to methods after pilot trial commencement (such as eligibility criteria), with reasons | 10, Table S1 |
| Participants | 4a | Eligibility criteria for participants | 5 |
| | 4b | Settings and locations where the data were collected | 7-8 |
| | 4c | How participants were identified and consented | 4,8 |
| Interventions | 5 | The interventions for each group with sufficient details to allow replication, including how and when they were actually administered | 6-7, Figure 1, published protocol |
| Outcomes | 6a | Completely defined prespecified assessments or measurements to address each pilot trial objective specified in 2b, including how and when they were assessed | Protocol |
| | 6b | Any changes to pilot trial assessments or measurements after the pilot trial commenced, with reasons | N/A |
| | 6c | If applicable, prespecified criteria used to judge whether, or how, to proceed with future definitive trial | 7, protocol |
| Sample size | 7a | Rationale for numbers in the pilot trial | 8 |
| | 7b | When applicable, explanation of any interim analyses and stopping guidelines | 8 |
| Randomisation: | | | |
| Sequence generation | 8a | Method used to generate the random allocation sequence | 6 |
| | 8b | Type of randomisation(s); details of any restriction (such as blocking and block size) | 6 |
| Allocation concealment | 9 | Mechanism used to implement the random allocation sequence (such as sequentially numbered containers), describing any steps taken to conceal the sequence until interventions were assigned | 6 |

| | | | |
|---|---|---|---|
| mechanism | | | |
| Implementation | 10 | Who generated the random allocation sequence, who enrolled participants, and who assigned participants to interventions | 6 |
| Blinding | 11a | If done, who was blinded after assignment to interventions (for example, participants, care providers, those assessing outcomes) and how | Protocol |
| | 11b | If relevant, description of the similarity of interventions | N/A |
| Statistical methods | 12 | Methods used to address each pilot trial objective whether qualitative or quantitative | 7-9 |
| **Results** | | | |
| Participant flow (a diagram is strongly recommended) | 13a | For each group, the numbers of participants who were approached and/or assessed for eligibility, randomly assigned, received intended treatment, and were assessed for each objective | 9, Figure 2 |
| | 13b | For each group, losses and exclusions after randomisation, together with reasons | 9, Figure 2 |
| Recruitment | 14a | Dates defining the periods of recruitment and follow-up | 9 |
| | 14b | Why the pilot trial ended or was stopped | 14 |
| Baseline data | 15 | A table showing baseline demographic and clinical characteristics for each group | Table 1 |
| Numbers analysed | 16 | For each objective, number of participants (denominator) included in each analysis. If relevant, these numbers should be by randomised group | Table 2, Table S2-S11 |
| Outcomes and estimation | 17 | For each objective, results including expressions of uncertainty (such as 95% confidence interval) for any estimates. If relevant, these results should be by randomised group | Table 2, Table S2-S11 |
| Ancillary analyses | 18 | Results of any other analyses performed that could be used to inform the future definitive trial | Figure 4 |
| Harms | 19 | All important harms or unintended effects in each group (for specific guidance see CONSORT for harms) | 11 |
| | 19a | If relevant, other important unintended consequences | N/A |
| **Discussion** | | | |
| Limitations | 20 | Pilot trial limitations, addressing sources of potential bias and remaining uncertainty about feasibility | 14 |
| Generalisability | 21 | Generalisability (applicability) of pilot trial methods and findings to future definitive trial and other studies | 12 |
| Interpretation | 22 | Interpretation consistent with pilot trial objectives and findings, balancing potential benefits and harms, and considering other relevant evidence | 12-15 |
| | 22a | Implications for progression from pilot to future definitive trial, including any proposed amendments | 13-15 |
| **Other information** | | | |
| Registration | 23 | Registration number for pilot trial and name of trial registry | 4 |
| Protocol | 24 | Where the pilot trial protocol can be accessed, if available | 4 |
| Funding | 25 | Sources of funding and other support (such as supply of drugs), role of funders | 28 |
| | 26 | Ethical approval or approval by research review committee, confirmed with reference number | 4 |

Citation: Eldridge SM, Chan CL, Campbell MJ, Bond CM, Hopewell S, Thabane L, et al. CONSORT 2010 statement: extension to randomised pilot and feasibility trials. BMJ. 2016;355. This is an Open Access article distributed in accordance with the terms of the Creative Commons Attribution (CC BY 3.0) license (http://creativecommons.org/licenses/by/3.0/), which permits others to distribute, remix, adapt and build upon this work, for commercial use, provided the original work is properly cited.

*We strongly recommend reading this statement in conjunction with the CONSORT 2010, extension to randomised pilot and feasibility trials, Explanation and Elaboration for important clarifications on all the items. If relevant, we also recommend reading CONSORT extensions for cluster randomised trials, non-inferiority and equivalence trials, non-pharmacological treatments, herbal interventions, and pragmatic trials. Additional extensions are forthcoming: for those and for up-to-date references relevant to this checklist, see www.consort-statement.org.

Appendix S2. Table 1 - Extended. Baseline Characteristics

| Baseline characteristic | Usual care* (n=27) | Fluid-sparing* (n=24) |
|---|---|---|
| Age (months) | 94.6 (67.9) | 118.9 (58.6) |
| Male Gender | 15 (55.6%) | 10 (41.7%) |
| Weight (kg) | 31.2 (27.4) | 33.9 (22.0) |
| Admission diagnosis to hospital | 27 (100%) | 24 (100%) |
| Sepsis-related | 25 (92.6%) | 22 (91.7%) |
| Not sepsis-related | 2 (7.4%) | 2 (8.3%) |
| Location of patient screened eligible | | |
| Emergency Department | 7 (25.9%) | 15 (62.5%) |
| Hospital Ward | 7 (25.9%) | 2 (8.3%) |
| PICU | 13 (48.1) | 7 (29.2%) |
| Previous medical comorbidities | 23 (85.2%) | 17 (70.8%) |
| Neurological | 10 (43.5%) | 11 (64.7%) |
| Cardiac | 5 (21.7%) | 2 (11.8%) |
| Pulmonary | 4 (17.4%) | 4 (23.5%) |
| Hematological | 5 (21.7%) | 8 (47.1%) |
| Malignancy | 5 (21.7%) | 4 (23.5%) |
| Gastrointestinal | 6 (26.1%) | 6 (35.3%) |
| Endocrine | 3 (13.0%) | 6 (35.3%) |
| Immunodeficiency | 1 (4.3%) | 1 (5.9%) |
| Genetic/Hereditary disorder | 8 (34.8%) | 6 (35.3%) |
| Renal | 5 (21.7%) | 4 (23.5%) |
| Other | 3 (11.1) | 5 (20.8) |
| PRISM III Score | | |
| Mean (SD) | 10.8 (7.6) | 14.5 (9.7) |
| Median (IQR) | 9.0 (5.0, 13.5) | 13 (7.5, 18.3) |
| Heart Rate (beats per minute) | 143.5 (26.7) | 134.3 (27.3) |
| Systolic Blood Pressure (mm Hg) | 91.8 (13.9) | 87.0 (14.6) |
| Diastolic Blood Pressure (mm Hg) | 53.4 (13.3) | 48.3 (9.5) |
| Mean Blood Pressure (mm Hg) | 67.0 (10.6); (n=14) | 59.7 (8.6); (n=11) |
| Capillary refill time (seconds) | 3 (2, 3); (n=20) | 3 (2, 4); (n=16) |
| Radial Pulse Quality | | |
| Normal | 18 (66.7%) | 10 (41.7%) |
| Weak or thready | 4 (14.8%) | 5 (20.8%) |
| Bounding | 2 (7.4%) | 2 (8.3%) |
| Not available (not documented) | 3 (11.1%) | 7 (29.2%) |
| Mental status altered from baseline | | |
| Yes | 18 (66.7%) | 17 (70.8%) |

| | | |
|---|---|---|
| Respiratory Rate (breaths per minute) | 30 (22, 40) | 22 (19.5, 31.5) |
| SpO2 (percent) | 97.0 (95.0, 98.5) | 98 (97, 99.3) |
| Body Temperature (degrees Celsius) | 37.6 (36.8, 38.4); (n=27) | 37.2 (36.9, 37.6); (n=23) |
| pH | 7.34 (7.25, 7.39); (n=23) | 7.32 (7.24, 7.41); (n=21) |
| Lactate (mmol/L) | 2.0 (1.5, 3.7); (n=21) | 2.9 (1.8, 4.0); (n=19) |
| Bicarbonate (mmol/L) | 22 (17, 24); (n=24) | 20 (16, 24); (n=22) |
| Glucose (mmol/L) | 6.6 (5.0, 7.3); (n=21) | 6.4 (4.8, 12.4); (n=19) |
| Potassium (mmol/L) | 3.9 (3.3, 4.3); (n=24) | 3.7 (3.4, 4.1); (n=21) |
| Documented positive test for Malaria | | |
| Not tested | 27 (100%) | 24 (100%) |

*estimates are mean (SD) or median (Q1, Q3) for continuous variables and n (%) for categorical variables.

Variable is available for group n unless otherwise noted.

Pediatric Intensive Care Unit (PICU); Pediatric Risk of Mortality (PRISM); standard deviation (SD); kilograms (kg); millimetre of mercury (mm Hg); interquartile range (IQR), oxygen saturation (SpO2)
